# Assessment of Willingness to Receive COVID-19 Vaccine and Associated Factors among Teachers in Dambi Dollo Town, Qellem Wallaga Zone, Oromia Regional State, Ethiopia: Institution based cross sectional study

**DOI:** 10.1101/2023.01.17.23284660

**Authors:** Desalegn Shiferaw, Chara Melaku, Lamessa Assefa, Tadele Kinati

## Abstract

**Background:** COVID-19 devastated the routine life of all human kind since its discovery in Wuhan, China in 2019 and caused by the severe acute respiratory syndrome coronavirus 2 (SARS-Cov-2) infections. Vaccination is an effective means for controlling the communicability of the disease and every effort has to be done to increase the proportion of vaccinated people against COVID-19.

**Objective:** The main objective of this study was to assess the willingness of teachers to receive COVID-19 vaccine and its associated factors in dambi dollo town, 2022.

**Methods:** School based cross sectional study design was applied. The data were collected self-administered questionnaire and analysed by SPSS version 23.0. Variables which showed association with dependent variable in the bivariate analyses at 0.25 were entered into multiple stepwise logistic regression model. P-Value 0.05 was considered statistically significant in this study. Adjusted Odds ratios together with corresponding 95% confidence intervals was used to interpret the findings.

**Results:** About 92% of the teachers in dambi dollo town have heard covid-19 vaccine and 67.2% of them know that the vaccine can prevent the COVID-19 disease. On the other hand 51 %(95%CI: 44.8, 57.2) of the respondents had good knowledge about COVID-19 vaccine. From the total 247 participants who have responded to our inquiry on their willing to receive the COVID-19 vaccine, 68.4% (95%CI: 62.5, 74.3) of them were willing to receive the vaccine immediately while the remaining were either not willing or not ready at the time of data collection. Those participants having good knowledge of the vaccine were about six times more willing to get vaccinated, (AOR=5.85, 95%CI: 2.74, 12.47) in comparison with those having poor knowledge of the vaccine.

**Conclusion:** In conclusion, the level of willingness to receive the COVID-19 Vaccine was 68.4% and relatively low in the current study population and participants’ religion and knowledge status are the two variables significantly associated with willingness to receive the vaccine.

## 1. Introduction

COVID-19 devastated the routine life of all human kind since its discovery in Wuhan, China in 2019 and caused by the severe acute respiratory syndrome coronavirus 2 (SARS-Cov-2) infections(1). Education sector is one of the seriously affected sectors by COVID-19 pandemic which hampered the normal teaching learning process at different levels.

To control spreading of coronavirus, countries have been taking different prevention strategies, such as social distancing, hand hygiene, partial and comprehensive lockdowns, closing schools and businesses, and/or wearing face masks in public(2,3) These measures are believed to be effective if strictly and consistently applied even though some of them are not applicable in long term especially in developing countries, where economy is already hit hard.

Vaccinations are a form of collective action, intended to produce herd immunity through mass participation in immunization programs which is the best option to prevent the severity of the disease and its spreading(4). However such immunity is not easily achieved unless a sufficient proportion of the population is vaccinated(5–7). Since COVID-19 was declared as the global pandemic, enormous numbers of companies and research centres around the world have been working hard to develop effective SARS-CoV-2 vaccines(8) and most of the are in action now.

National regulatory authorities have granted use of several COVID-19 vaccines. Some of these have been approved for emergency or full use by at least one WHO-recognized stringent regulatory authority (Oxford–AstraZeneca, Pfizer-BioNTech, Sinopharm-BBIBP, Moderna, Sinovac, and Johnson & Johnson) (9).

The global effort through COVAX and the donation from different countries and organizations is not a guarantee for receiving of the vaccine in order to avoid the severe form of the disease and meet the target of vaccination(10). Different studies have revealed that the acceptance of the vaccine among different segments of the community in different countries is not uniform and facing challenges of hesitance and less willingness to receive(11). A study conducted in Mozambique showed that only 64.9% of general population were willing to accept the COVID-19 vaccination (12). Another study conducted in Kenya among school teachers revealed that less than thirty eight per cent of the participants declared their readiness to accept the vaccine if it is safe (13). A similar study conducted in northeast Ethiopia among university students showed that 69.3% of them have showed willingness to accept the vaccine (14) while school teachers in gonder town of Ethiopia indicated that only 54.8% of all the study participant showed above the median on intension to receive the vaccine (15). The findings of the above mentioned studies point that still there are assignments to do among the community especially among the high risk groups like school teachers if control of COVID-19 has to be improved significantly. Beyond this, such lower level acceptance for the vaccine puts countries at risk of not reaching their target and leads to the wastage of the scarce amount of the vaccine given to them. Therefore, this study aimed to identify the extent of the willingness to receive the vaccine among the school teachers especially in our current study area since there is no such study conducted there and to act upon the identified factors for the low acceptance of the vaccine.

## 2. Methods and materials

### 2.1. Study area and period

This study was conducted in dambi dollo town, Qellem wallaga zone of Oromia regional state; Qellem Wallaga zone is among the 20 zones of the region located in western Ethiopia. The capital of this zone, Dambi Dollo, is located at about 637 km to the southwest of Addis Ababa the capital city of Ethiopia. Based on the 2007 Census conducted by the <u>Central Statistical Agency</u> of Ethiopia (CSA), this Zone has a total population of 997,666, of whom 401,905 are men and 395,761 women. While 76, 277 or 9.56% are urban inhabitants. Dambi dollo town is with five high schools and six primary schools. The study was conducted from November 2021 to February 2022.

### 2.2. Study Design

Institution based cross sectional study design was applied.

### 2.3. Population

#### 2.3.1. Source population

The source population was all teachers in primary and secondary school teachers and Dambi Dollo University teachers, which is found in dambi dollo town, Qellem wallaga zone.

#### 2.3.2. Study population

The study population was teachers in the selected schools and departments during this study.

### 2.4. Eligibility Criteria

#### 2.4.1. Inclusion criteria

Teachers who were on their job and able to provide information during the data collection period were included in this study.

#### 2.4.2. Exclusion criteria

Those teachers who were critically ill and couldn’t provide necessary information were out of this study.

### 2.5. Sample size determination

The required sample size was calculated using single population proportion formula

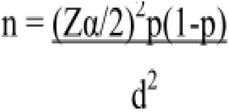

Where; n= sample size, considering the following assumptions;

P= proportion of school teachers willing to accept the COVID-19 vaccination, which was 54.8% from the study conducted in Gonder town, Ethiopia(15).

Zα/2= 95% confidence level which was (1.96),

d=5% marginal error and 10% non-response rate, and by using correction formula finally resulted in total sample size of 267.

### 2.6. Sampling procedure

Dambi dollo town was purposely selected based on its proximity and accessibility in the current contest of the zone. This is because of security instability in our current study area which didn’t allow to go out of the capital of the zone.

Firstly, stratification was done based on ownership of the schools into private and governmental. Then, some governmental institutions including dambi dollo University and some private schools were selected using simple random sampling technique and the calculated sample size was proportionally allocated. Finally, the study participants from each school were selected randomly using lottery method.

### 2.7. Variables and measurements

#### 2.7.1. Outcome Variable

The outcome variable in this study was willingness to take COVID-19 vaccine which was measured through (Yes/No) question. For the sake of regression analysis those who responded not decided were assumed as No and analysed.

#### 2.7.2. Independent Variables

- Socio-demographic factors: these include age, sex, level of education, place of work, level of teaching, religion, and monthly salary and whether they have known chronic disease.
- COVID-19 vaccine related knowledge: some of the questions included under this category are have you ever heard of COVID-19 vaccine, source of information, COVID-19 can be prevented by vaccine, do you know any vaccine against COVID-19, the vaccine is for free and voluntarily.
- COVID-19 vaccine related attitudes. These variables were whether they are at higher risk, essential for them, the vaccine is safe, the disease can be prevented by the vaccine, and the vaccine is against culture and religion.

### 2.8. Operational Definitions

COVID-19 vaccine acceptance was measured using “Yes” and “No” questions. Respondents were asked “Are you willing to be vaccinated against COVID-19?” Ten items were used for assessing respondents’ knowledge about the COVID-19 vaccine. Those who correctly answered the question were given one point, while incorrect responses were given zero values. Respondents who have scored >=70% were assumed as having good knowledge while those who scored less than 70% were considered as having poor knowledge towards the COVID-19 vaccine.

Eleven items were used to assess the respondents’ attitude on the COVID-19 vaccine. Those who agreed received 3 points, neutral 2 points, and disagreed got 1 point for positive questions and vice versa for negative quoted attitude questions. The respondents’ attitude ranged from 11 to 33, with a cut-off of greater than or equal to 70% (23–33) were considered as a positive attitude while less than 70% (23) were taken as having a negative attitude towards the COVID-19 vaccine.

### 2.9. Data collection tools and procedures

The data were collected using pretested, structured, and self-administered questionnaire developed by the investigators from relevant and similar literatures(10,16,17). First, the tool was prepared in English language and then translated into the local language, Afan Oromo. Pre-test was done on 5% of the total sample size and necessary modifications were done accordingly.

Six public health professionals were assigned to collect the data and another 3 senior public health professionals have supervised data collectors in the process of data collections and the investigators have supervised the overall activity.

### 2.10. Data quality control

To assure the quality of data, the following measures were undertaken including pre-testing of the questionnaire. The final version of the questionnaire was translated into the local language of the respondents (Afan Oromo), and two days of intensive training was given to data collectors and strict supervision was provided.

### 2.11. Data processing and analysis

Data were first checked manually for completeness and then coded; entered, cleaned using Epidata version 4.6.0.6 and exported to SPSS version 23.0 for analysis. Descriptive analysis was used to describe the percentages and number of distributions of the respondents by socio-demographic and other characteristics. The main Statistical method applied was logistic regression, both the bivariate and multivariate analyses. Variables which showed association with the outcome variable in the bivariate analyses at 0.25 were entered into multiple logistic regression model. P-Value 0.05 was considered statistically significant in this study. Adjusted Odds ratios together with corresponding 95% confidence interval was used to interpret the findings.

### 2.12. Ethical considerations

Official Letter was secured from Dambi Dollo University Research and Technology Innovative directorate office. Having the official letter from the university, it was presented to Qellem wallaga education department and finally to each school. Written consent was obtained from the participants and assuring them about their right to participate or not and continued with those study participants agreed to participate. On the other hand research ethical approval letter was obtained from dambi Dollo University research and technology innovation directorate (the authorised office to provide the ethical clearance in the University). Confidentiality and privacy was maintained by excluding the name and ID of study participants from the questionnaire. Those who were not volunteer to participate in the study were respected not to participate by their decision.

## Results

### 2.13. Socio-demographic characteristics

A total of 267 questionnaires were distributed among teachers in dambi dollo town teaching at different levels of schooling with a total response rate of 92.51%. From the total study participants responded, 46.2% were within 30-39 years age range and 72% of them were male. About 61% were degree holders and 89.5% were teaching in public schools among which 52.6% were high school (9-12) teachers (Table 1).

**Table 1.**
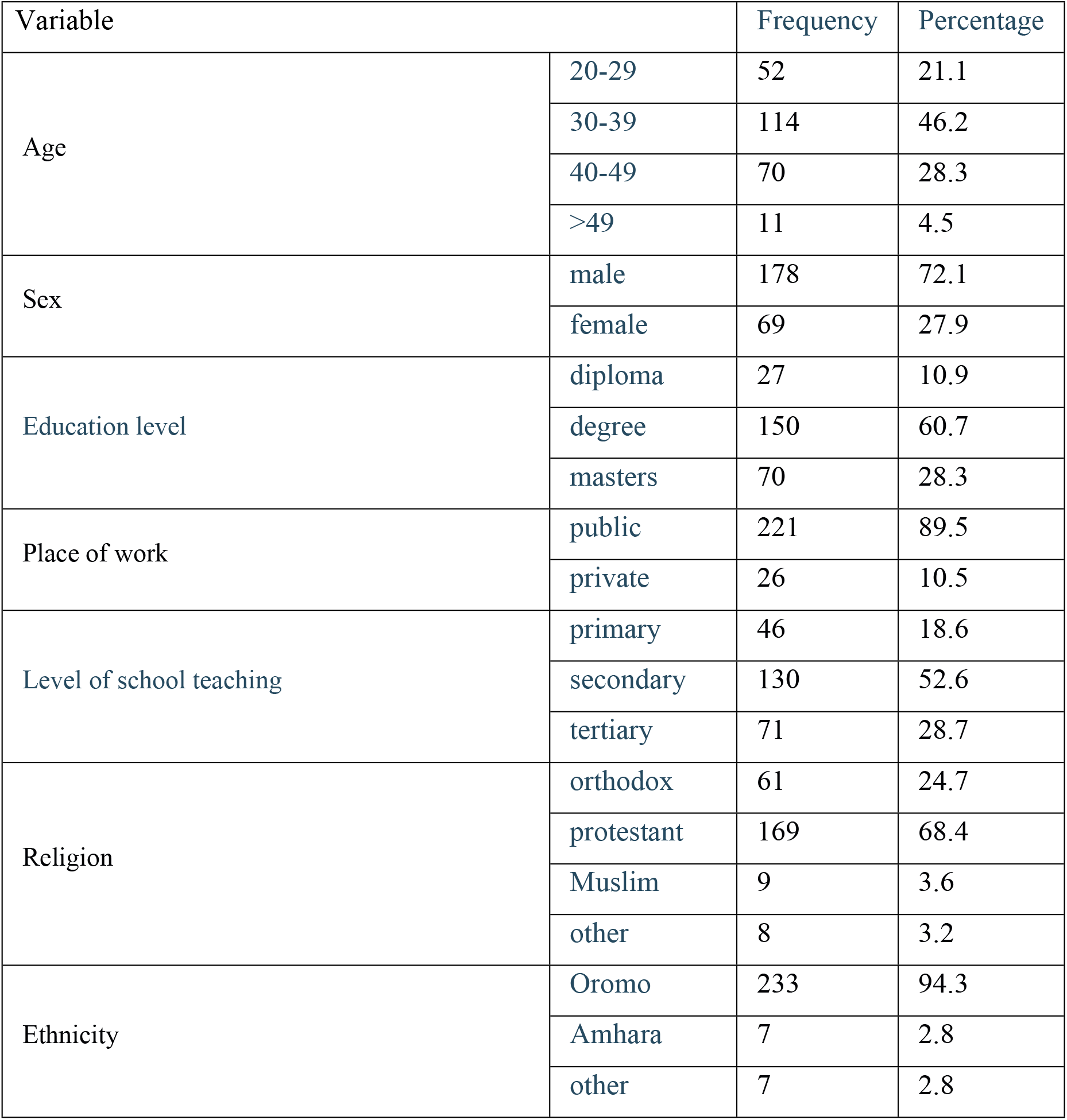
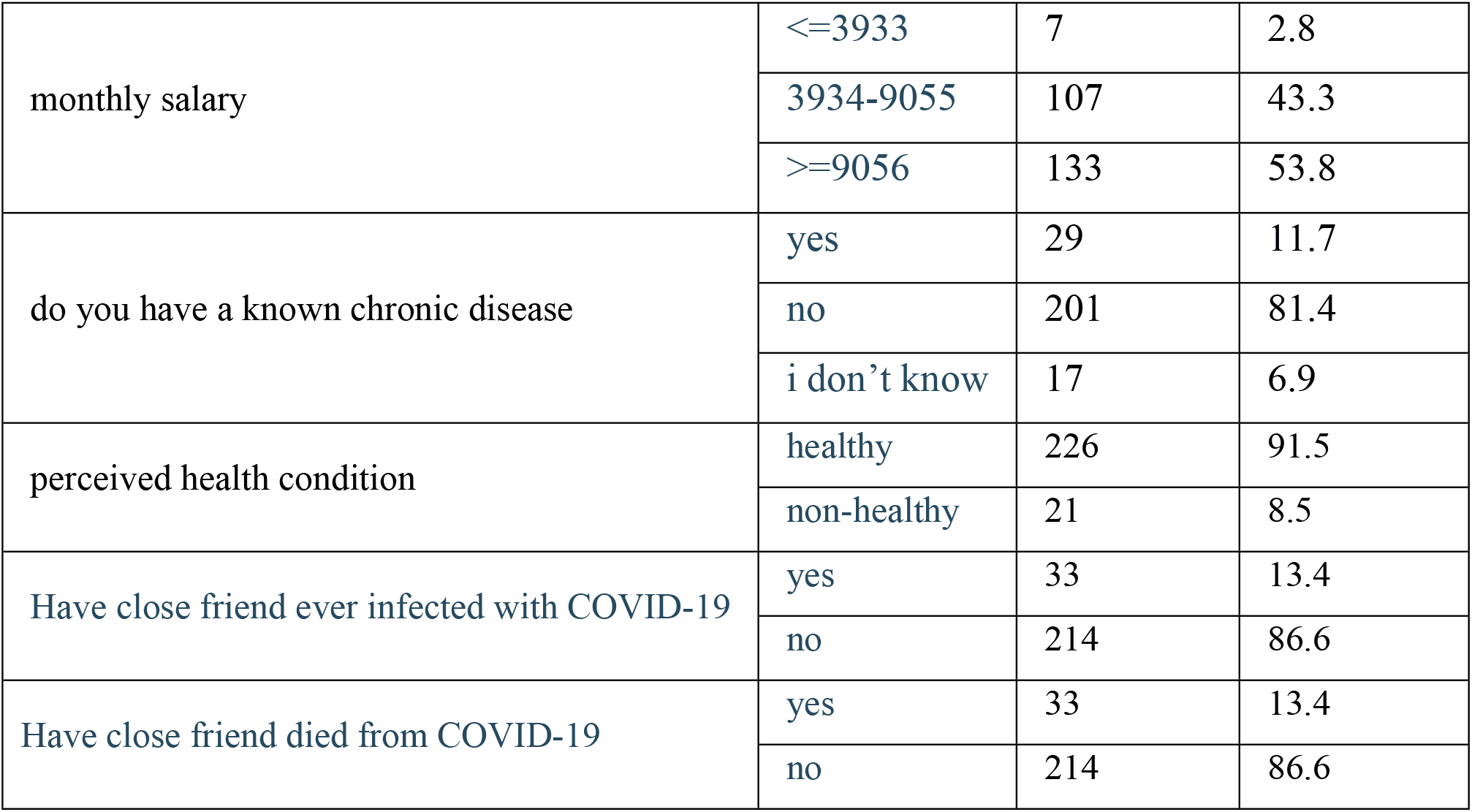
Socio-demographic characteristics of the study participants on assessment of willingness to receive COVID-19 Vaccine and its associated factors among teachers in Dambi dollo town, Qellem Wallaga Zone, Oromia regional state, Ethiopia, 2022.

### 2.14. Knowledge about COVID-19 vaccine among teachers in dambi dollo town

In this study, about 92% of the respondents have heard about the COVID-19 vaccine and more than half of them mentioned that TV was their source of information. About 67% of them said that the vaccine saves from the disease and 77.7% of them know that the provision of the vaccine is voluntarily. In general, only 51% of the participants were having a good knowledge regarding the COVID-19 vaccine (Table 2).

**Table 2.** Knowledge of the participants towards receiving COVID-19 Vaccine and its associated factors among teachers in Dambi dollo town, Qellem Wallaga Zone, Oromia region, Ethiopia, 2022.

| Variables |  | Frequency | Percentage (%) |
| --- | --- | --- | --- |
| Have you ever heard about COVID-19 vaccine | yes | 227 | 91.9 |
|  | no | 20 | 8.1 |
| From whom have you heard about covid-19 vaccine | radio | 37 | 15.0 |
|  | TV | 132 | 53.4 |
|  | social media | 59 | 23.9 |
|  | health professionals | 14 | 5.7 |
| COVID-19 can be prevented by the vaccine | yes | 166 | 67.2 |
|  | No | 17 | 6.9 |
|  | I don't know | 64 | 25.9 |
| Which of the following COVID-19 vaccine have you heard | AstraZeneca | 93 | 37.7 |
|  | Johnson and Johnson | 53 | 21.5 |
|  | BioNTech, pfizer | 5 | 2.0 |
|  | sinopharm | 4 | 1.6 |
|  | I don't know | 92 | 37.2 |
| The vaccine is given two times within 28 days apart | yes | 40 | 16.2 |
|  | No | 32 | 13.0 |
|  | i don't know | 175 | 70.9 |
| The vaccine is provided for free in Ethiopia | yes | 171 | 69.2 |
|  | No | 11 | 4.5 |
|  | I don't know | 65 | 26.3 |
| The provision of the vaccine is based on voluntary not obligatory | yes | 192 | 77.7 |
|  | No | 15 | 6.1 |
|  | I don't know | 40 | 16.2 |
| Healthcare professionals, chronic patients, elders and school teachers are the prioritized groups for vaccination | yes | 165 | 66.8 |
|  | No | 39 | 15.8 |
|  | I don't know | 43 | 17.4 |
| The vaccine of COVID-19 has started in dambi dollo town | yes | 148 | 59.9 |
|  | No | 27 | 10.9 |
|  | I don't know | 72 | 29.1 |
| In Ethiopia currently COVID-19 vaccine is available for all >12 years | yes | 59 | 23.9 |
|  | No | 75 | 30.4 |
|  | I don't know | 113 | 45.7 |
|  | Total | 247 | 100.0 |

### 2.15. Willingness to receive covid-19 vaccine

The main variable intended to assess in this study was the willingness of the school teachers to receive the vaccine. Accordingly, 68.4% of the participants responded that they are willing to receive the COVID-19 vaccine.

Who need to take COVID-19 Vaccine?

The other point assessed was for whom the vaccine is needed and the study participants have reflected their understanding. Accordingly, about 56.3% of them responded that the vaccine is needed for everyone while 8.1% of them even don’t know for whom the vaccine is intended.

#### Who need to take the vaccine?

While we were assessing participants’ perceptions towards for whom the vaccine is important, 56.3% of them responded that everyone needs to take the COVID-19 vaccine (fig1).

**Fig. 1.**
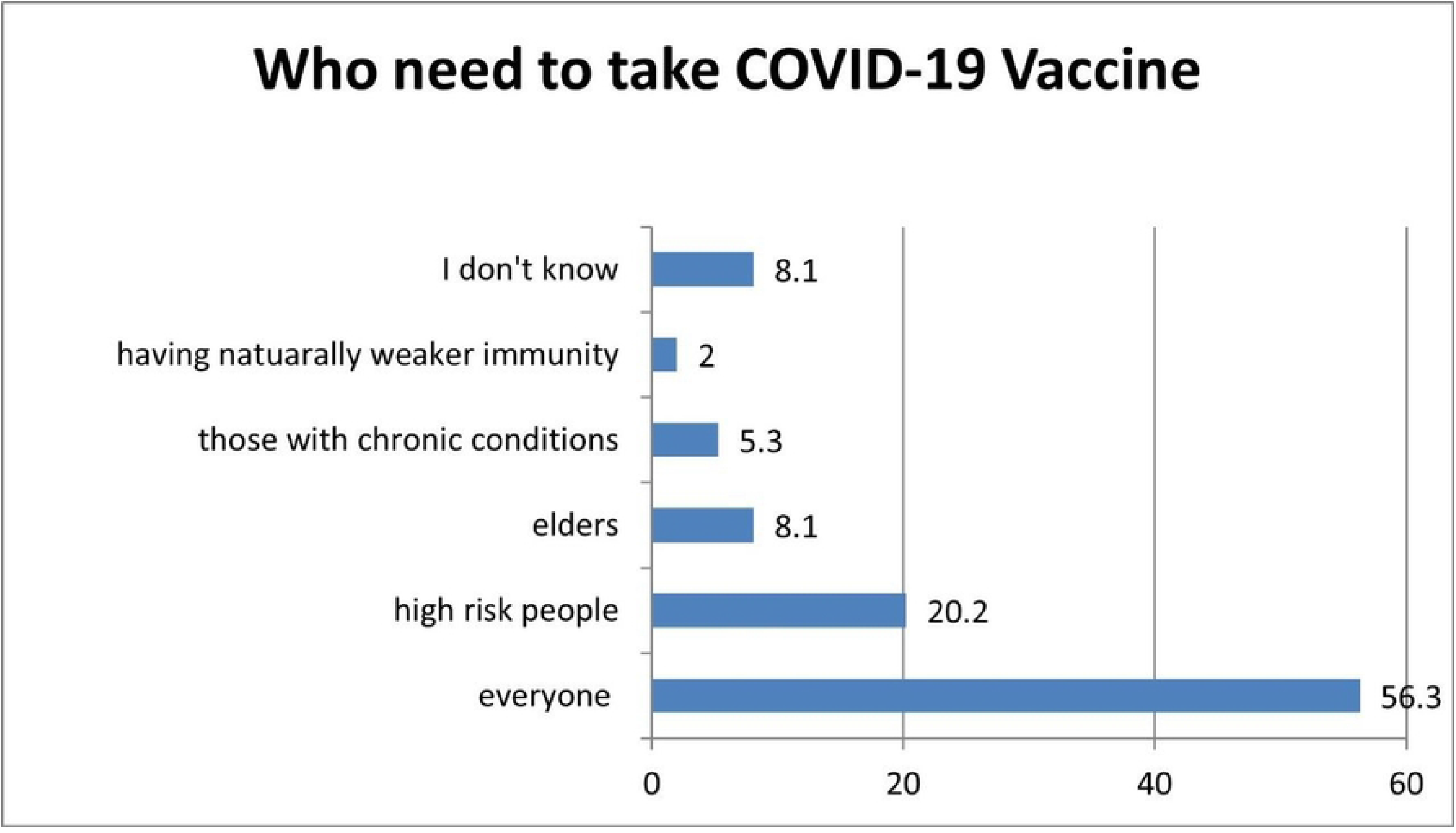
Respondents’ perception towards who need to receive COVID-19 vaccine in dambi dollo, 2022.

### 2.16. Attitude towards COVID-19 vaccine among teachers in dambi dollo town

According to the finding of this study, 61.5% of the participants believe that they were at higher risk of the infection and 64% of them believed that the vaccine is essential for them. About 55% of the respondents believed that the vaccine has to be given to all while 50.6% of them believed that the provision of the vaccine is not against their culture. Generally, only 28.3% of the participants had good attitude towards receiving the COVID-19 vaccine (Table 3).

**Table 3.** Attitude of respondents toward COVID-19 vaccine in dambi dollo town, 2022.

| Variable |  | Frequency | Percentage (%) |
| --- | --- | --- | --- |
| Do you believe that you are at a higher risk of being infected by COVID-19? | Agree | 152 | 61.5 |
|  | Neutral | 38 | 15.4 |
|  | Disagree | 56 | 23.1 |
| Is the COVID-19 vaccine essential for you? | Agree | 158 | 64.0 |
|  | Neutral | 43 | 17.4 |
|  | disagree | 46 | 18.6 |
| Do you believe that COVID-19 Vaccine is safe for you? | agree | 140 | 56.7 |
|  | neutral | 60 | 24.3 |
|  | disagree | 47 | 19.0 |
| Do you think COVID-19 can be prevented by the vaccine? | agree | 133 | 53.8 |
|  | neutral | 75 | 30.4 |
|  | disagree | 39 | 15.8 |
| The vaccine of covid-19 should be given to all | agree | 136 | 55.1 |
|  | neutral | 60 | 24.3 |
|  | disagree | 51 | 20.6 |
| COVID-19 vaccine is against your culture | agree | 41 | 16.6 |
|  | neutral | 81 | 32.8 |
|  | disagree | 125 | 50.6 |
| COVID-19 vaccine is against your religion | agree | 56 | 22.7 |
|  | neutral | 79 | 32.0 |
|  | disagree | 112 | 45.3 |
| Do you think that you are susceptible to COVID-19 | agree | 114 | 46.2 |
|  | neutral | 70 | 28.3 |
|  | disagree | 63 | 25.5 |
| It is not possible to reduce the incidence of COVID-19 without vaccination | agree | 49 | 19.8 |
|  | neutral | 75 | 30.4 |
|  | disagree | 123 | 49.8 |
| Do you think that the vaccine makes you infected by COVID-19 | agree | 36 | 14.6 |
|  | neutral | 114 | 46.2 |
|  | disagree | 97 | 39.3 |
| I believe that the side effects of the vaccine outweigh its immunizing advantages | agree | 49 | 19.8 |
|  | neutral | 85 | 34.4 |
|  | disagree | 113 | 45.7 |

### 2.17. Associated factors of willingness to receive COVID-19 vaccine among the respondents

As evidenced from the output of the bivariate analyses, few variables have shown association with the willingness to receive the vaccine while others were not. Accordingly, all variables with P-value <0.25 were seen under multivariable logistic regression to if they have significant association with the dependent variable as shown in table below.

According to the bivariate analysis output, the current level the teachers were teaching, religion, having chronic disease, having close friend died of the disease, knowledge of the vaccine and attitude towards the vaccine had an association whether the teachers were willing to receive the vaccine. But, after adjustment for confounding situation only two variables remained having statistically significant association with the outcome variable. Based on this, religion and knowledge status of the study participants were significantly associated variables with the outcome variable. Accordingly, protestant religion followers were less likely to receive the by 63.2%, (AOR=0.368, 95%CI: 0.161, 0.840) while Muslim were less likely to receive the vaccine by 92.2%, (AOR=0.078, 95%CI: 0.010, 0.580) compared with Orthodox Christian followers. On the other hand, school teachers with good knowledge regarding the COVID-19 vaccine were times more likely to receive the vaccine, (AOR=5.85, 95%CI: 2.744, 12.473) compared with those with poor knowledge (Table 4).

**Table 4.** Association between socio-demographic variables and willingness to receive COVID-19 vaccine among teachers in dambi dollo town (n=247) in Qellem Wallaga Zone, Oromia, Ethiopia, 2022.

| Variable | Number | Willing to receive the vaccine (%) | COR(95% CI) | AOR(95% CI) |
| --- | --- | --- | --- | --- |
| Highest Educational level attained |  |  |  |  |
| Diploma | 27 | 18(66.7) | 1 | 1 |
| Degree | 150 | 93(62.0) | 0.816(.043,1.938) | 1.31(0.381,4.501) |
| Masters | 70 | 58(82.9) | 2.42(0.877,6.657) | 2.481(0.574,10.720) |
| Level of school currently teaching |  |  |  |  |
| Primary | 46 | 35(76.1) | 1 | 1 |
| Secondary | 130 | 76(58.5) | 0.442(0.206,0.498) | 0.455(0.152,1.362) |
| Tertiary | 71 | 58(81.7) | 1.402(0.567,3.469) | 0.586(0.165,2.086) |
| Religion |  |  |  |  |
| Orthodox | 61 | 49(80.3) | 1 | 1 |
| Protestant | 169 | 110(65.1) | 0.457(0.225,0.925) | 0.368(0.161,0.840) |
| Muslim | 9 | 3(33.3) | 0.122(0.027,0.561) | 0.078(0.010,0.580) |
| Other | 8 | 7(87.5) | 1.714(0.192,15.292) | 0.965(0.088,10.591) |
| Do you have chronic disease |  |  |  |  |
| Yes | 29 | 24(82.8) | 4.267(1.101,16.537) | 4.524(0.825,24.820) |
| No | 201 | 136(67.7) | 1.86(0.686,5.041) | 1.331(0.392,4.523) |
| I don't know | 17 | 9(52.9) | 1 | 1 |
| perceived health condition |  |  |  |  |
| Healthy | 226 | 157(69.5) | 1.707(0.687,4.237) | 1.427(0.366,5.557) |
| Non-healthy | 21 | 12(57.1) | 1 | 1 |
| Do you have a close relative or friend ever infected by COVID-19? |  |  |  |  |
| Yes | 33 | 25(75.8) | 1.519(0.652,3.539) | 1.253(0.439,3.575) |
| No | 214 | 144(67.3) | 1 | 1 |
| Do you have a close relative or friend died of COVID-19? |  |  |  |  |
| Yes | 33 | 29(87.9) | 3.832(1.298,11.315) | 3.379(0.90, 12.696) |
| No | 214 | 140(65.4) | 1 | 1 |
| Knowledge status |  |  |  |  |
| Good | 126 | 110(87.30) | 7.225(3.832,13.621) | 5.85(2.744, 12.473) |
| poor | 121 | 59(48.76) | 1 | 1 |
| Attitude status |  |  |  |  |
| Good | 70 | 62(88.57) | 5.07(2.288,11.234) | 0.572(0.216,1.514) |
| Bad | 177 | 107(60.45) | 1 | 1 |

## 3. Discussion

This study was conducted to assess the willingness of teachers to receive the COVID-19 vaccine in dambi dollo town. It has included teachers from primary school, high school and college/university and 247 teachers have responded with 92.5% response rate.

The struggle of controlling the pandemic of COVID-19 will not bear satisfactory outcome without universal access and utilization of the COVID-19 vaccine. Specific to developing countries like Ethiopia, unfavourable level of willingness to take the vaccine triangulated with the difficulty of accessing required doses of the vaccine is feared to allow the pandemic to intensify the multi-sectorial impact in such countries. Unlike other similar studies, our current study included teachers from different levels of schooling including primary, secondary and higher education institution to see if there is differences among the different levels which is helpful for intervention.

According to this study, about 92% of the teachers in dambi dollo town have heard covid-19 vaccine and 67.2% of them know that the vaccine can prevent the COVID-19 disease. In this study, 51 %(95%CI: 44.8, 57.2) of the respondents had good knowledge about COVID-19 vaccine. The finding of our study was lower than that of studies conducted among college students in Gonder town, 69.2%(18) and health professionals in Wallaga University referral hospital, 66%(19). Regarding their attitudes towards COVID-19 vaccine, only 28.3% of the participants of our current study had overall positive attitude. This finding was lower compared with that of the study conducted among college students of gonder town, 51% (18). This difference might be due to the fact that college students are more flexible to accept changes and ideas as they are younger ages and very close to new developments as a result of their engagement with numerous media from where they get rich information.

From the total 247 participants who have responded to our inquiry whether they were willing to receive the COVID-19 vaccine, 68.4% (95%CI: 62.5, 74.3) of them were willing to receive the vaccine immediately while the remaining were either not willing or not ready at the time of data collection. The level of willingness to receive the COVID-19 vaccine among teachers at different levels in dambi dollo town was comparable with the results of studies conducted in Ghana, 70 %(20), among university students in northern Ethiopia, 69.3 %(21) and finding of research conducted among health professionals of Wallaga University referral hospital, 65.4%(19). On the other, the willingness to receive COVID-19 vaccine among the participants of the current study was lower than that of the study conducted in central and southern Italy undergraduate students, which was 90.1% (22). The level of willingness to be vaccinated among our participant was higher than the finding of research conducted in dessie hospital hospital, 59.4%(23), research conducted in southern Ethiopia, 46.1%(24), among residents of Sodo town, 45.5%(17), among college students of Gonder town, 34.2%(25) and among residents of south western Ethiopia, 29.8%(26). These differences might be attributed to different reasons as the vaccine is relatively new. Understanding among different areas and different segments of community is different and in turn it can affect their attitude regarding the vaccine.

From all the variables checked for having association with the outcome variable, those with P-value less than 0.25 on bivariate analysis were entered to multivariable analysis to see if they have significant association. Accordingly, nine independent variables were enrolled for multivariable analysis from which only two variables, namely religion of the respondent and their knowledge about the vaccine were remained significantly associated with the outcome variable.

Based on this, protestant Christians were less willing to receive the vaccine by about 63%, (AOR=0.37, 95%CI: 0.168, 0.840) and Muslim were less willing by 92%, (AOR=0.078, 95%CI: 0.010, 0.580) compared with the Orthodox Christian followers in our current study. This finding indicated that there is a need for awareness creation by the leaders of the respective religion towards the vaccine. On the other hand, those participants having good knowledge of the vaccine were about six times more willing to get vaccinated, (AOR=5.85, 95%CI:2.744,12.473) in comparison with those having poor knowledge of the vaccine. The presence of significant association between vaccine knowledge and the willingness to receive it reiterates the importance of capitalizing on the gained knowledge and increasing its coverage through different means.

## 4. Limitations of the study

Like most survey studies, our current study may have different limitations. One of these limitations is the way we categorize the good and poor knowledge may not clearly differentiate their understanding. Individuals with a knowledge of >=70 may not be willing to receive the vaccine and vice versa. The other limitation is that we have included teachers from three different levels primary, secondary and higher education institution, who might be with different level of understanding about the disease and the vaccine. On the other hand our current study area is far from the centre of the country and having frequent network interruption which might affect their access to timely and adequate information regarding the vaccine which can affect their willingness.

## 5. Conclusion

In conclusion, the level of willingness to receive the COVID-19 Vaccine was 68.4% among teachers of different levels in dambi dollo town which is not enough to control the pandemic. In this study, participants religion and knowledge status are the two variables significantly associated with willingness to receive the vaccine. On the other hand, this figure is alarming as the participants were teachers expected to be role model for others and all concerned bodies should take seriously and put extra effort to promote the vaccine acceptance level.

## Data Availability

additional files will be provided by the authors upon request

## 6. Acknowledgment

We would like to thank Dambi Dollo University for facilitating the whole activities of the study and encouraging us in all means. We are so much grateful to all who participated in the process of conducting the study including our respondents, data collectors and friends.

## 9. Supporting Information

S1 Fig 1. Respondents’ perception towards who need to receive COVID-19 vaccine in dambi dollo, 2022 (in MS word)

S1 Fig 1. Respondents’ perception towards who need to receive COVID-19 vaccine in dambi dollo, 2022 (in tiff image)

S1 Table 1. Socio-demographic characteristics of the study participants on assessment of willingness to receive COVID-19 Vaccine and its associated factors among teachers in Dambi dollo town, Qellem Wallaga Zone, Oromia regional state, Ethiopia, 2022.

S2 Table 2. Knowledge of the participants towards receiving COVID-19 Vaccine and its associated factors among teachers in Dambi dollo town, Qellem Wallaga Zone, Oromia region, Ethiopia, 2022.

S3 Table 3. Attitude of respondents toward COVID-19 vaccine in dambi dollo town, 2022.

S4 Table 4. Association between socio-demographic variables and willingness to receive COVID-19 vaccine among teachers in dambi dollo town (n=247) in Qellem Wallaga Zone, Oromia, Ethiopia, 2022.

S1 questionnaire. Willingness to receive COVID-19 vaccine assessment tool

